# Risk of reactivation of hepatitis B virus (HBV) and tuberculosis (TB) and complications of hepatitis C virus (HCV) following Tocilizumab therapy: A systematic review to inform risk assessment in the COVID era

**DOI:** 10.1101/2021.03.22.21254128

**Authors:** Cori Campbell, Monique Andersson, M Azim Ansari, Olivia Moswela, Siraj A Misbah, Paul Klenerman, Philippa C Matthews

## Abstract

Tocilizumab (TCZ), an IL-6 receptor antagonist, is used in the treatment of COVID. However, this agent carries a ‘black box’ warning for infection complications, which may include reactivation of tuberculosis (TB) or hepatitis B virus (HBV), or worsening of hepatitis C virus (HCV). Due to the pace of clinical research during the COVID pandemic, prospective evaluation of these risks has not been possible. We undertook a systematic review, generating mean cumulative incidence estimates for reactivation of HBV and TB at 3.3% and 4.3%. We could not generate estimates for HCV. These data derive from heterogeneous studies pre-dating the COVID outbreak, with differing epidemiology and varied approaches to screening and prophylaxis. We underline the need for careful individual risk assessment prior to TCZ prescription, and present an algorithm for clinical stratification. There is an urgent need for ongoing collation of safety data as TCZ therapy is used in COVID.

**KEY POINTS:** Use of tocilizumab treatment in COVID-19 may risk infective complications. We have undertaken a systematic literature review to assess the risks of reactivation of HBV and TB, generating mean estimates of 3.3% and 4.3% incidence, respectively.

## INTRODUCTION

IL-6 is an immunomodulatory cytokine, performing diverse roles in homeostasis and in the outcomes of infectious, inflammatory and autoimmune disease (Figure 1A) [1]. Elevated levels are associated with immune response to infection [2], and also with metabolic and cardiovascular risk [3]. Tocilizumab (TCZ) is a ‘biologic’ agent, a recombinant humanized IgG1 monoclonal antibody that competitively blocks both the soluble and membrane-bound forms of the interleukin-6 (IL-6) receptor (Figure 1B). Licensed for the treatment of rheumatoid arthritis, juvenile idiopathic arthritis, giant cell arteritis [4,5], and for the management of a cytokine storm caused by CAR-T therapy, it is also used off-label for the treatment of other refractory autoimmune or inflammatory disease.

**Figure 1:**
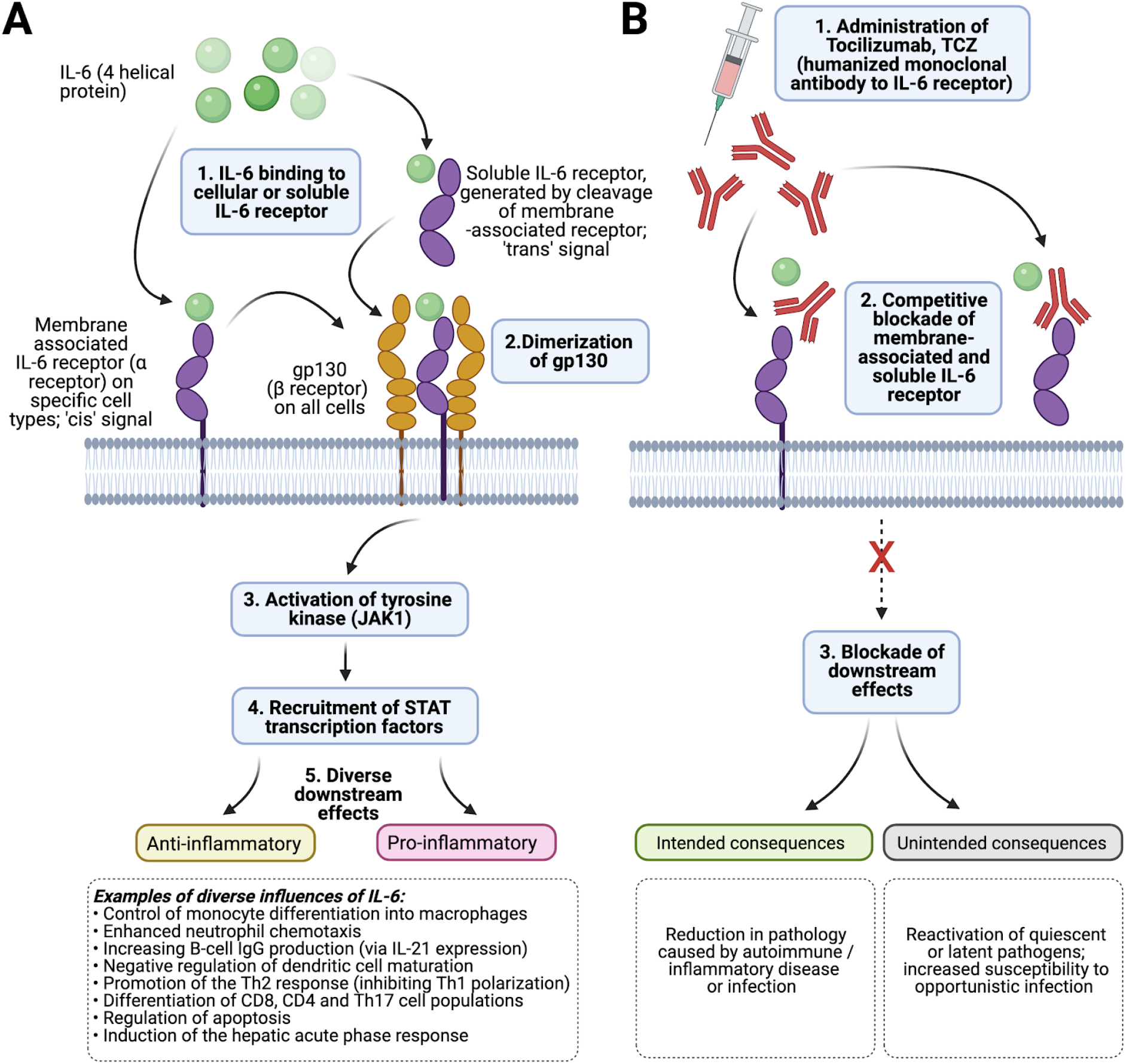
Diagram to show (A) the mechanism of action of IL-6 and (B) the influence of tocilizumab, TCZ. The influence of IL-6 in viral infection is reviewed by Rose-John *et al*., [1] *and Velazquez-Salinas et al*. [2]. gp130 is expressed on all cells in the body, IL-6 receptor is expressed by specific cells only (hepatocytes, monocytes/macrophages, CD4+ T cells, some epithelial cells). Cis-(‘classical’) signalling is mediated through cell-associated IL-6 receptor, and associated with housekeeping, while trans-signalling describes the pathway associated with free soluble IL-6 receptor which may be more relevant in response to infection. JAK-1 -Janus Kinase 1. STAT -signal transducer and activator of transcription. Figure created with Biorender.com, with licence to publish.

**Figure 2:**
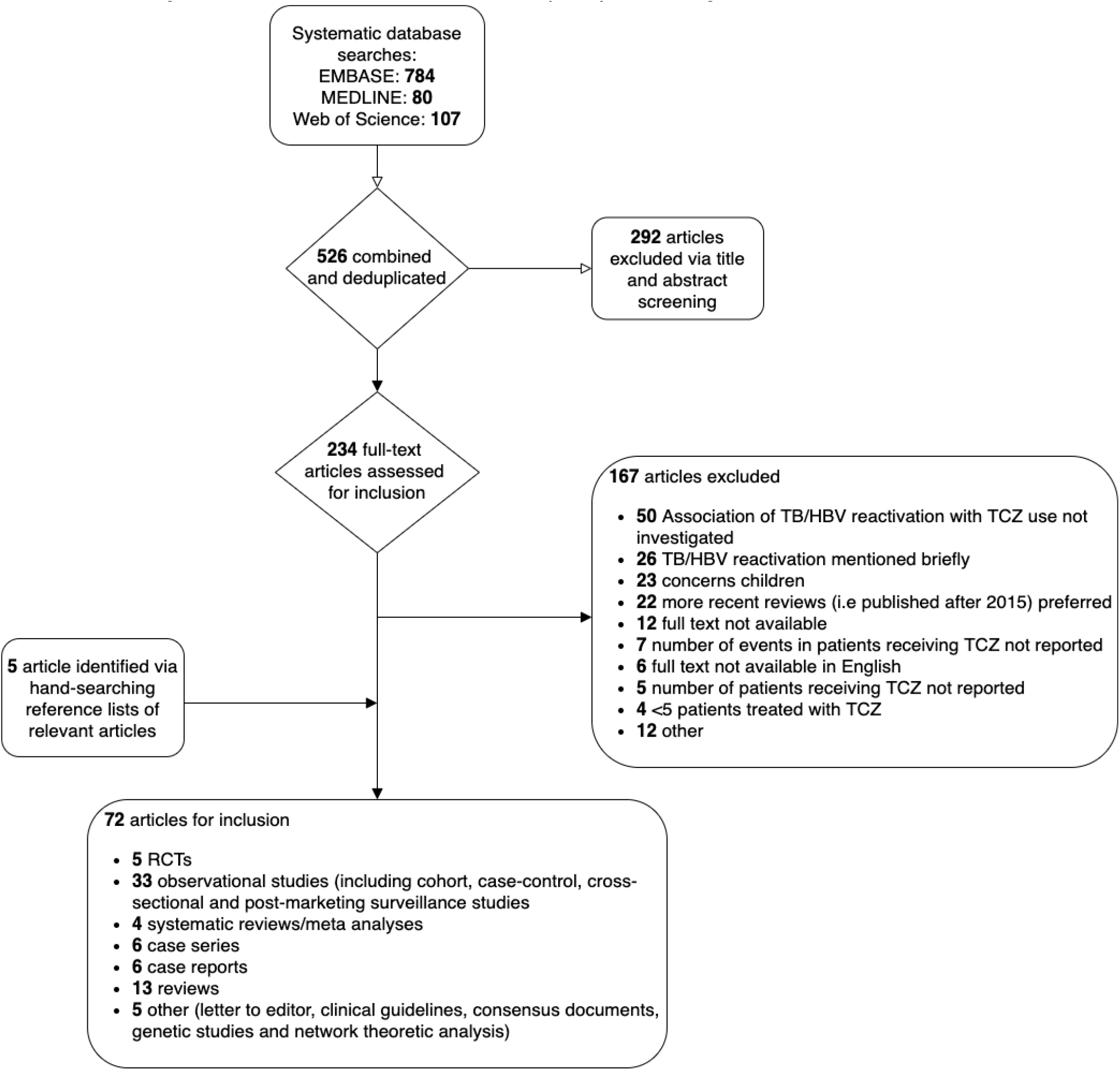
Flow chart to show selection of studies reporting the risk of HB and/or TB reactivation in patients treated with tocilizumab (TCZ) from a systematic literature review.

COVID-19, caused by infection with SARS-CoV-2, can be characterised by a ‘cytokine storm’ associated with a severe systemic inflammatory response syndrome, immune dysregulation and a pro-thrombotic state. Elevated IL-6 levels have been associated with the need for invasive mechanical ventilation in these patients [6]. For this reason, TCZ has appeal as a therapeutic agent, and multiple studies have set out to determine its role in COVID patients. The literature is currently heterogeneous [7]; several clinical studies have reported a benefit (e.g. [8–11]), or find modest improvements in length of hospital stay without reducing deaths [12], while others do not find significantly improved outcomes [13] or even identify possible harm [14]. Meta-analysis undertaken in January 2021 which included 71 studies (of which 6 were randomised trials) reported overall improved survival associated with TCZ therapy (adjusted mortality risk HR 0.52) [15].

Expert recommendations suggest consideration of a single dose of TCZ (8mg/kg of actual body weight, up to a maximum of 800mg) in individuals with respiratory failure secondary to COVID-19 admitted to a critical care environment [16], or with signs of severe or critical disease and elevated markers of systemic inflammation [17]. TCZ is recommended in addition to dexamethasone, which has been widely adopted as standard of care based on findings from the RECOVERY trial [18]. TCZ half life is dose-dependent and estimated at 8-14 days [19]; some recommendations suggest a repeat dose after 12–24 hours if there has not been sufficient clinical improvement [20]. Due to small overall numbers of COVID patients treated, limited prospective follow-up, and lack of concordance in reported clinical outcomes, guidance recognises the low certainty of the evidence and the pressing need for more data [7,21].

To date, the safety profile of TCZ in COVID patients is deemed good [7,13], albeit based on small numbers, delivery of treatment only in selected clinical settings, and limited periods of follow-up. However, based on experience from the pre-COVID era, TCZ carries a black box warning for serious infection events (SIEs; Figure 1B) caused by a range of opportunistic bacterial, mycobacterial, viral and fungal pathogens that may lead to hospitalisation or death. Pooled analyses of the SIE risk associated with the standard dose have estimated incidence at ∼5 per 100 patient years [22,23], similar to other biologic drugs [1], but with higher risk in older adults, and when combined with steroids and/or methotrexate [24–26], or with previous biologic therapy [27].

TB and Hepatitis B virus (HBV) reactivation are of particular concern due to the high global burden (Table 1). Expert opinion and guidelines, pre-dating the COVID pandemic, recommend for screening for both TB and HBV infection before starting TCZ, to inform risk stratification, surveillance, and treatment or prophylaxis [25,26,28,29] (Table 1). Data are limited, as patients who screened positive for hepatitis were excluded from pre-marketing authorisation TCZ clinical trials. Furthermore, large clinical COVID studies using TCZ therapy have not yet been conducted in settings where TB and/or HBV prevalence are high, and without sufficient length of follow up to quantify the risk of reactivation of these infections, or to determine which individuals are at highest risk. HCV infection is another blood-borne virus for which outcomes may be complicated by treatment with biologic agents, although this pathogen does not have the potential for latency, and outcomes are thought to be less severe than those of HBV in patients receiving biologic therapies [26].

**Table 1:**
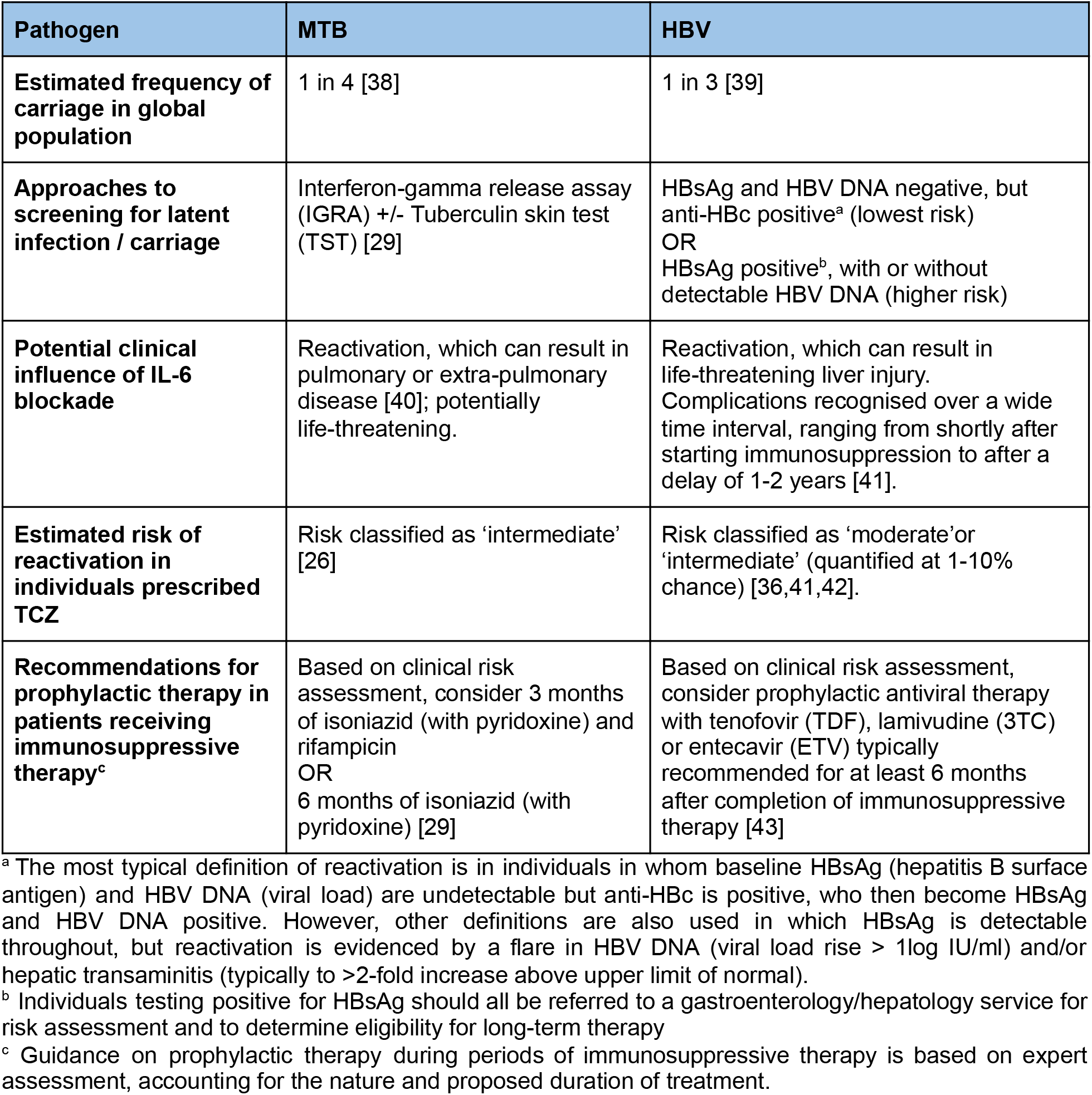
Features of Mycobacterium tuberculosis (MTB) and Hepatitis B Virus (HBV) infection pertinent to the use of therapeutic IL-6 blockade.

As more patients are prescribed TCZ for COVID, there is an urgent need for data to inform risk stratification and management of COVID patients at elevated risk of these infective complications. We therefore set out to collate data for HBV and TB reactivation, and for complications of active HCV infection, in the context of TCZ therapy, to consider how this evidence may apply to COVID patients and to highlight where further action is needed.

## METHODS

### Literature search

We searched the global WHO database of Individual Case Safety Reports (ICSRs) / adverse drug reactions (ADRs) (‘VigiBase’) on 21st February 2021 [30]; search terms are presented in Table S1. We undertook a systematic literature review (on 25 February 2021), searching Medline, Embase and Web of Science databases using search terms presented in Table S2, in accordance with PRISMA guidelines (Table S3). We searched all databases from 1 January 2002 to 25 February 2021. No further search restrictions were applied. We included any article (including abstracts) reporting/investigating the risk of adverse HBV, TB and or hepatitis C virus (HCV) outcome(s) associated with TCZ use. Results from each database were combined and deduplicated prior to eligibility screening. Reference lists of relevant systematic reviews/meta-analyses were also searched to identify studies for inclusion.

### Data extraction and statistical analysis

Studies were considered eligible for inclusion if they were observational cohort or post-marketing surveillance studies reporting i) the number of patients in the study sample receiving TCZ and ii) the number of TCZ-treated patients experiencing an outcome of interest (TB or HBV reactivation). For each study, we recorded country, publication year, study design, study population, follow-up period, number of participants receiving TCZ, number of TB/HBV reactivation cases, age at baseline (mean/median), and sex. We also collated information pertaining to screening of participants for TB/HBV at baseline, as this is a potential source of bias.

We calculated mean cumulative incidence for HBV and TB reactivations from observational cohort studies. Individual and mean cumulative incidence estimates were displayed in forest plots constructed in R (version 4.0.2) using the ‘metafor’ package (version 2.4-0 [https://cran.r-project.org/web/packages/metafor/metafor.pdf]). Upper 95% confidence intervals were calculated for studies reporting zero events of interest using the rule of three [31].

## RESULTS

### ‘Vigibase’ WHO database reports HBV and TB reactivation in patients prescribed TCZ

The VigiBase database [30] holds 47,205 records for complications of TCZ, of which 14,147 (30%) pertain to ‘infections and infestations’, with tuberculosis mentioned in 135 cases (0.9% of infections), and HBV in 42 (0.3% of infections) (Table S1). However, further interpretation and risk quantification is not possible based on this source, as the database does not contain any denominator data (total number of individuals treated with TCZ therapy), no details of clinical or demographic history, nor the indication/dose/duration of TCZ therapy.

### Systematic review to determine the risk of TB reactivation in patients receiving TCZ therapy

We identified 19 observational cohort studies in which the incidence of TB reactivation was reported, with a follow up duration of between 24 weeks and 11 years (Figure 3A; Table S4). Studies were typically small, with a median size of 49 participants, although six studies reported on data for >1000 individuals. The incidence of TB reactivation varied from no reported cases (in six studies; Table S4), up to 31% (but with wide confidence intervals, 95% CI 5.4, 66.1) in an Indian cohort [32]. This variation is likely to reflect significant differences in risk between settings according to the population prevalence of latent TB, as well as varied approaches to screening and prophylaxis. Although the Indian study reports risk assessment and prophylaxis, the incidence of reactivation was nevertheless high [32]. In contrast, a study in South Africa -another high prevalence setting -recorded only 0.8% incidence of TB reactivation (95% CI 0, 7.2) [33], which is likely to reflect successful chemoprophylaxis. Several studies did not report their approach to screening and prophylaxis (Table S4), thus making direct comparisons between studies difficult.

**Figure 3:**
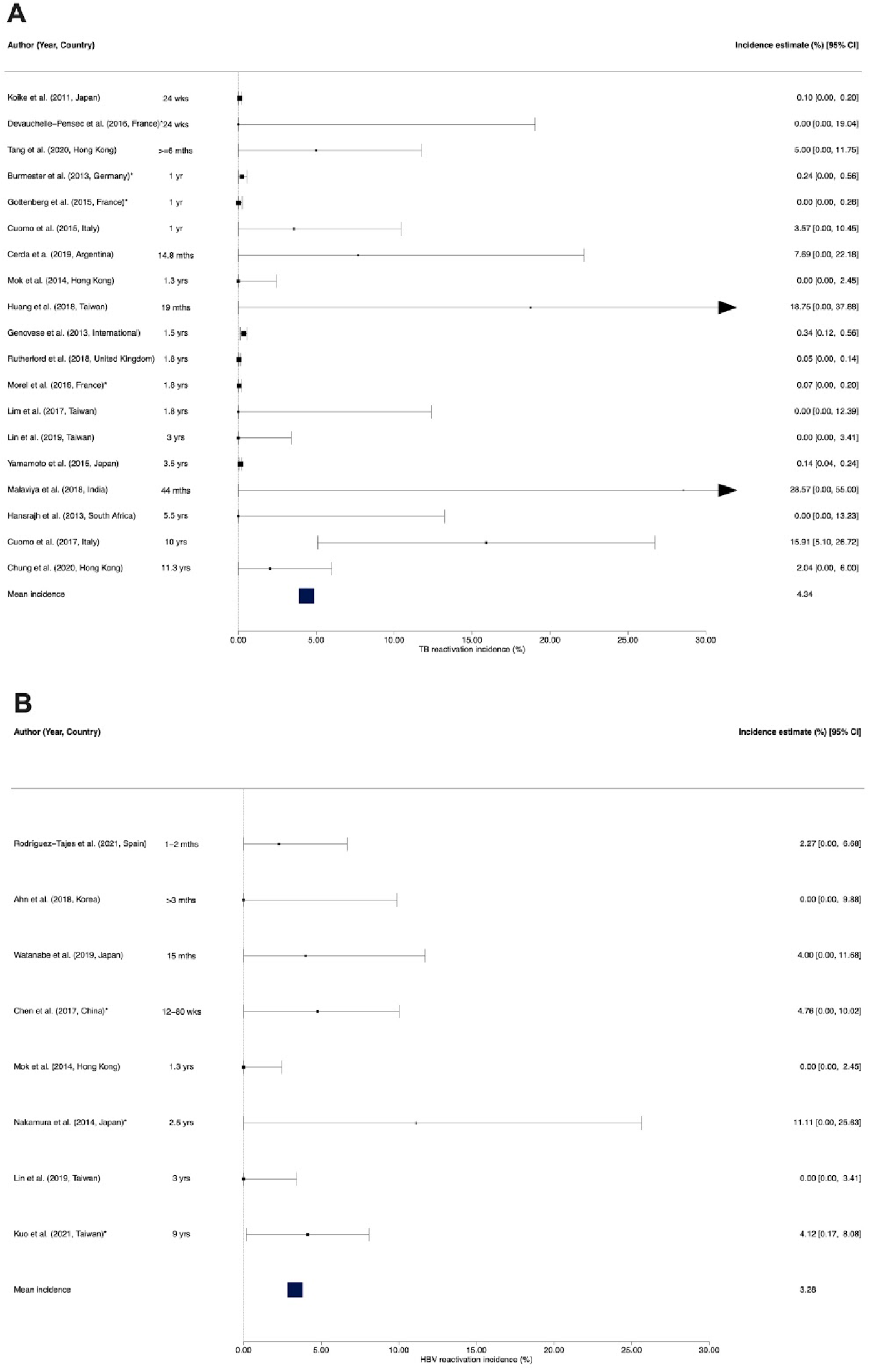
Forest plots to show data from a systematic literature review to identify studies reporting a risk of reactivation of (A) TB and (B) HBV in individuals treated with TCZ therapy. Studies are ranked in order of follow-up duration. Location of the study is presented for each study, as the baseline prevalence of TB and HBV infection varies substantially by geography. Reactivation incidence (x-axis) shows proportion of events among all those treated over the time period observed. The size of each point estimate is proportional to cohort size. A pooled mean estimate is presented, but should be interpreted with caution due to the difficulties in combining heterogeneous data from diverse settings.

A mean cumulative incidence estimate from the available data puts the estimated risk of TB reactivation at 4.3% at a population level, but this figure should be interpreted with caution on the basis of the heterogeneity described above.

### Systematic review to determine the risk of HBV reactivation in patients receiving TCZ therapy

We identified 8 observational cohort studies in which the risk of HBV reactivation was reported, with a median of 46 study participants (range 18-159); (Figure 3B; Table S5). Follow-up was undertaken over periods ranging from 1 month up to 9 years, with no cases of HBV reactivation reported in three cohorts, but up to as high as 13% in a Japanese cohort [34]. Baseline prevalence of HBsAg and anti-HBc varied between cohorts, and approaches to screening and prophylaxis were undertaken differently between settings, with a more rigorous approach in some centres that may explain the lower incidence of reactivation events.

A mean cumulative incidence estimate from the available data puts the estimated risk of HBV reactivation at 3.3% at a population level, but as for TB these figures should be interpreted with caution and individual risk assessment is key.

### Systematic review to determine the risk of complications of HCV infection in patients receiving TCZ therapy

We identified six studies which investigated or discussed risk of HCV complications associated with TCZ use (Table S6). Most studies reported low risks of HCV flare associated with TCZ use, but nevertheless called for future investigations and screening of rheumatoid arthritis patients before administration of anti-rheumatic treatment. The scarcity of evidence in this field has been previously highlighted [35]

### Review articles

Review and guidance articles are unified in highlighting potential risks of HBV, HCV and TB infection, although with varying risk appraisal (Table S7). Most regard the risk of HBV or TB reactivation associated with TCZ therapy as ‘moderate’ or ‘intermediate’. Typically, screening is recommended for HBV and TB (±HCV) prior to starting any biologic therapy, and prophylaxis is recommended in those deemed to be at the highest risk of complications (summarised in Figure 4).

**Figure 4:**
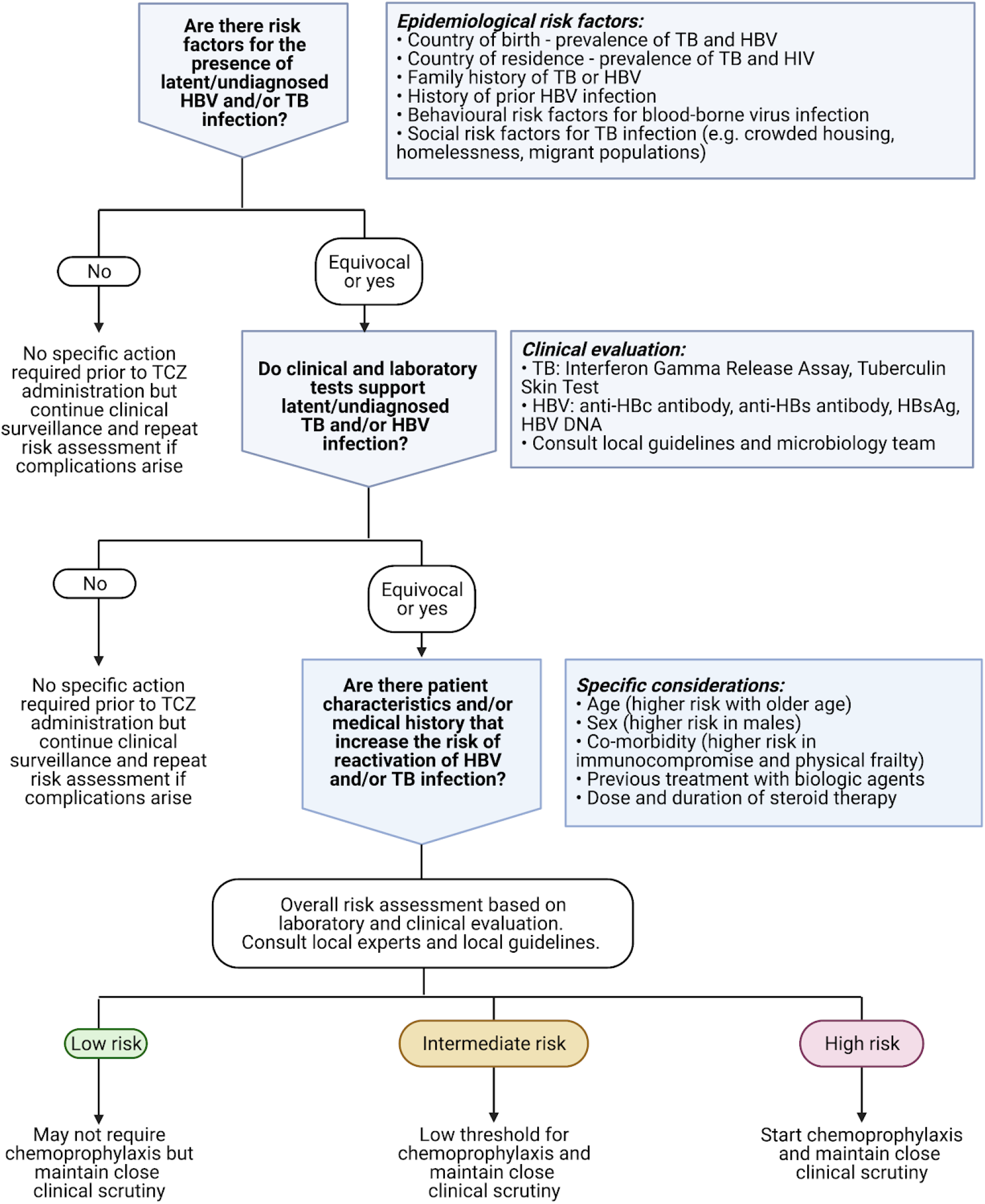
Algorithm to underpin clinical approach to risk assessment for latent or undiagnosed TB or HBV prior to commencing treatment with TCZ. Existing recommendations for anti-cytokine therapy suggest considering antiviral prophylaxis especially in patients who are older in age, male sex, frail, and/or have underlying haematological malignancy [36]. Figure created with Biorender.com, with licence to publish.

### Risk of bias

Characteristics of the individual studies may have contributed to bias. Populations studied may be skewed against reactivation events (if studies are not conducted in high risk populations, or if individual patients at high risk are given prophylaxis or actively excluded from trial participation). Conversely other studies set out to enrich for individuals with prior HBV exposure such that risks for a general population may be over-estimated. Examples of each of these sources of bias are presented with details in Tables S4 (for TB) and S5 (for HBV). Durations of follow-up were short in most studies, likely making them insufficient to detect all possible TB or HBV reactivation events. Baseline approach to screening and/or administration of prophylaxis for TB and/or HBV was not always reported, and was inconsistent across studies.

## DISCUSSION

### Summary/new insights

TCZ is gaining traction as therapy for patients at the severe end of the clinical COVID-19 spectrum, and has been incorporated into clinical recommendations [16,17], which will result in its more widespread use in a critically ill patient group at risk of infective complications because of the global prevalence of latent TB and HBV carriage (Table 1). Our summary data demonstrate a small risk of HBV and TB reactivation overall, with estimates of 3.3% and 4.3% respectively for a population treated with TCZ. However, our ability to quantify these risks is constrained due to limited and heterogeneous data. The risk at a population level may have been systematically underestimated to date, due to bias resulting from the implementation of screening and prophylaxis, low risk populations enrolled in clinical studies, and insufficient durations of follow-up. The risk for individuals with the latent HBV and TB infections is likely much higher, as the population estimates include individuals that do not carry latent infections. We highlight the imperative for individualised risk assessment in patients prescribed TCZ therapy, as the incidence of infective complications may be substantially higher in certain at-risk groups.

### Specific clinical recommendations

Consensus from existing guidelines recommend screening for carriage of HBV and TB prior to TCZ therapy. However, even where recommendations are established, screening and prophylaxis are often not consistently undertaken [28,36], and more data are needed to better inform risk assessment and prescription of prophylactic therapy. Chemoprophylactic agents are widely available, can be administered enterally, are cheap, safe, effective and well-tolerated, so these interventions generally carry a low risk. In individuals with COVID at high epidemiological risk of these infections, our data suggest that there should be a low threshold for screening, and risk-assessment for prophylaxis (Figure 4). However, it is important to consider the real world practicalities of screening in different settings, and to avoid mandates for screening that cause unintended delays in therapy. For HBV, serum markers (HBsAg, anti-HBc) are generally accessible and affordable, with short turn-around times. For TB screening, Quantiferon and Mantoux may not be immediately available, or may take longer to organise in practice, leading to delays before a result is available.

Recommendations are relevant not just at the point of TCZ prescription but during follow-up, with clinical awareness of HBV and TB as potential complications in the weeks and months after TCZ administration. As TCZ may blunt markers of infection (such as CRP), reactivation events can be more difficult to diagnose in this group [1]. In individuals with potential risk of HBV reactivation, monitoring of liver enzymes and HBV markers (HBsAg and viral load), should be considered.

In the case of HCV, flares are possible on biologic therapy [ref], and as successful direct acting antiviral (DAA) treatment can now be prescribed, screening is advocated in anyone with risk factors for infection and/or with unexplained abnormalities in liver function tests, such that therapy can be started.

### Caveats

Estimates of risk may be biased, with a likelihood of under-estimating true risk based on selection of low-risk study participants, provision of prophylaxis when indicated, short durations of follow-up and reporting population estimates which include individuals without latent infection. We were unable to undertake a formal meta-analysis due to the heterogeneity of data, and the reported mean across studies must be interpreted with caution. Further ‘real world’ data are needed to investigate the impact of TCZ in racially diverse COVID cohorts, in patients with other underlying pathology, and over longer periods of follow-up[1,16], both to determine the subgroups in whom treatment may be of most benefit, and to identify the circumstances in which it carries the most risk. It is notable that many existing publications about the use of TCZ in COVID do not even discuss the potential for these infective complications.

Risk appraisal for use of TCZ in COVID is currently based on experience in treating patients with inflammatory and rheumatological disease, who may also be immunosuppressed as a result of their primary condition, and in whom immunosuppressive therapy is prolonged and may involve multiple agents (including steroids, methotrexate and other biological agents). However, although COVID treatment comprises only one or two doses of TCZ, the context of critical illness, universal combination with steroid therapy, and two doses given in quick succession, may inflate the risks. On all of these grounds, extrapolation of existing TCZ risk assessment data to a population with COVID infection must be undertaken with caution. As the field of COVID therapeutics is expanded, there is interest in using other immunomodulatory drugs [37], which may also be associated with an increase in infective complications.

We have elected to exclude data on children, due to a current focus on TCZ use to manage severe COVID, which arises almost exclusively in adults. However, this should not preclude the collection of robust safety data for TCZ use in children for rheumatological disease. We recognise that due to our exclusion criteria we may have overlooked some paediatric data that could be relevant to the use of this agent in adults.

From existing data, we are unable to determine the highest risk period for SIEs with respect to the timing of TCZ dosing, and it is possible that more TB and HBV reactivation events would be identified by longer periods of follow-up, although this trend is not clear from the small number of studies we have identified. However, it is most likely that the greatest risk in COVID therapy is within a short number of weeks of the TCZ dose. Similarly, there are currently no data to determine whether the risk is inflated in individuals receiving two doses rather than one dose of TCZ.

### Future questions

There is a need for wider data and more robust guidelines to support the safe use of biologic therapy with appropriate risk assessment and consistent use of antimicrobial prophylaxis, equitable inclusion of diverse populations in research and clinical studies, and ongoing prospective surveillance of patients receiving TCZ to identify complications. In the longer term, better biomarkers that will improve risk stratification, improving the identification of patients at the highest risk of HBV reactivation who stand to gain most from prophylaxis.

In addition to the pathogens we have assessed here, there are other SIEs associated with TCZ therapy that remain to be addressed in individuals with COVID, including invasive fungal infection, atypical mycobacterial infection, herpes viruses (among which VZV reactivation may be a specific concern), and other opportunistic infections. More data are needed to inform screening, risk assessment, prophylaxis and treatment of these infections in COVID patients. Given that these events are typically uncommon, there is a crucial need for experience to be collated and centralised. The WHO ‘Vigibase’ platform provides a potential foundation for such activity, but collection of enhanced metadata will be required to determine the individuals at risk of SIEs.

### Conclusions

Clinicians prescribing TCZ for patients with COVID should be aware of infective complications, have a low threshold for screening for HBV and TB, consider prophylaxis in those with identified risk factors, and maintain surveillance over the weeks and months following therapy. There is an urgent need for more data in diverse populations in order to inform improved approaches to risk assessment and prevention of these potentially serious complications.

## Data Availability

All data presented were extracted from published articles identified via systematic review.

## ACKNOWLEDGEMENTS

Nil

## SUPPL MATERIAL: provided as a separate file

**TABLE S1:** Search terms and studies identified through a search of WHO ‘VigiBase’ database.

**TABLE S2:** Search terms used in systematic search of Embase, MEDLINE and Web of Science databases.

**TABLE S3:** PRISMA checklist

**TABLE S4:** Summary characteristics of observational studies investigating risk of TB infection in individuals receiving tocilizumab.

**TABLE S5:** Summary characteristics of observational studies investigating risk of HBV infection in individuals receiving tocilizumab.

**TABLE S6**: Summary characteristics of studies investigating risk of HCV infection in individuals receiving tocilizumab.

**TABLE S7**: Summary of review articles presenting data on the risks of HBV, HCV and/or TB reactivation in patients on TCZ therapy, identified by systematic literature review

