## Supplementary materials for "Risk of reactivation of hepatitis B virus (HBV) and tuberculosis (TB) and complications of hepatitis C virus (HCV) following Tocilizumab therapy: A systematic review to inform risk assessment in the COVID era"

**CONTENTS:**

**TABLE S1:** Search terms and studies identified through a search of WHO 'VigiBase' database.

**TABLE S2:** Search terms used in systematic search of Embase, MEDLINE and Web of Science databases.

**TABLE S3:** PRISMA checklist

**TABLE S4:** Summary characteristics of observational studies investigating risk of TB infection in individuals receiving tocilizumab.

**TABLE S5:** Summary characteristics of observational studies investigating risk of HBV infection in individuals receiving tocilizumab.

**TABLE S6**: Summary characteristics of studies investigating risk of HCV infection in individuals receiving tocilizumab.

**TABLE S7**: Summary of review articles presenting data on the risks of HBV, HCV and/or TB reactivation in patients on tocilizumab therapy, identified by systematic literature review

**Table S1: Results from a search of 'Vigibase', WHO database of Individual Case Safety Reports (ICSRs) / adverse drug reactions (ADRs) ('VigiBase') (https://www.who-umc.org/vigibase/vigibase/) for reports of TB or HBV complications in the context of tocilizumab (TCZ) therapy.** Search conducted on 21-Feb-2021. The database reports a total of 47205 records for TCZ, among which 14147 (30%) pertain to 'infections and infestations'.

| Pathogen | Complication reported | Number reported |
|---|---|---|
| Tuberculosis | Tuberculosis | 85 |
| | Pulmonary tuberculosis | 29 |
| | Latent tuberculosis[a] | 5 |
| | Peritoneal tuberculosis | 4 |
| | Disseminated tuberculosis | 3 |
| | Lymph node tuberculosis | 3 |
| | Pericarditis tuberculosis | 2 |
| | Bone tuberculosis | 1 |
| | Joint tuberculosis | 1 |
| | Extra-pulmonary tuberculosis | 1 |
| | Tuberculosis pleurisy | 1 |
| | **Total TB complications (% of all reported infection complications)** | **135 (0.9%)** |
| Hepatitis B virus | Hepatitis B reactivation | 25 |
| | Hepatitis B | 12 |
| | Viral hepatitis[b] | 4 |
| | Acute hepatitis B | 1 |
| | **Total HBV complications (% of all reported infection complications)** | **42 (0.3%)** |

[a] Latent tuberculosis listed as a complication, but uncertain whether this reflects an episode of reactivation resulting in clinical disease.
[b] Viral hepatitis listed without an aetiological agent; uncertain whether this category includes HBV or other causes of viral hepatitis.

**Table S2**: Search terms used in systematic search of Embase, MEDLINE and Web of Science databases

| Criteria | Database | Search terms |
|---|---|---|
| Population | MEDLINE and Embase (searched via Ovid) | (HBV or HCV or "hepatitis B virus" or "hepatitis infection" or "hepatitis C virus" or tuberculosis or TB or "Mycobacterium tuberculosis").af. |
| | Web of Science | TS=(HBV or HCV or "hepatitis B virus" or "hepatitis infection" or "hepatitis C virus" or tuberculosis or TB or "Mycobacterium tuberculosis") |
| Exposure | MEDLINE and Embase (searched via Ovid) | (tocilizumab or atlizumab or "interleukin-6 receptor").af. |
| | Web of Science | TS=(tocilizumab or atlizumab or "interleukin-6 receptor*") |
| Outcome | MEDLINE and Embase (searched via Ovid) | (infection or reactiv* or exacerbat* or worse*)).ab. |
| | Web of Science | TS=(infection or reactiv* or exacerbat* or worse*) |
| Final search | MEDLINE and Embase (searched via Ovid) | ((HBV or HCV or "hepatitis B virus" or "hepatitis infection" or "hepatitis C virus" or tuberculosis or TB or "Mycobacterium tuberculosis").af.) AND ((tocilizumab or atlizumab or "interleukin-6 receptor").af.) AND ((infection or reactiv* or exacerbat* or worse*)).af.) |
| | Web of Science | (TS=(HBV or HCV or "hepatitis B virus" or "hepatitis infection" or "hepatitis C virus" or tuberculosis or TB or "Mycobacterium tuberculosis")) AND (TS=( tocilizumab or atlizumab or "interleukin-6 receptor*" )) AND (TS=(infection or reactiv* or exacerbat* or worse*)) |

MH, medical subject heading term.

**Table S3: PRISMA checklist for a systematic review** [https://doi.org/10.1136/bmj.b2700]

| Section/topic | # | Checklist item | Reported in section |
|---|---|---|---|
| **TITLE** | | | |
| Title | 1 | Identify the report as a systematic review, meta-analysis, or both. | Abstract |
| **ABSTRACT** | | | |
| Structured summary | 2 | Provide a structured summary including, as applicable: background; objectives; data sources; study eligibility criteria, participants, and interventions; study appraisal and synthesis methods; results; limitations; conclusions and implications of key findings; systematic review registration number. | Unstructured abstract requested by journal. |
| **INTRODUCTION** | | | |
| Rationale | 3 | Describe the rationale for the review in the context of what is already known. | Included in introduction |
| Objectives | 4 | Provide an explicit statement of questions being addressed with reference to participants, interventions, comparisons, outcomes, and study design (PICOS). | Included in introduction |
| **METHODS** | | | |
| Protocol and registration | 5 | Indicate if a review protocol exists, if and where it can be accessed (e.g., Web address), and, if available, provide registration information including registration number. | No protocol or registration number |
| Eligibility criteria | 6 | Specify study characteristics (e.g., PICOS, length of follow-up) and report characteristics (e.g., years considered, language, publication status) used as criteria for eligibility, giving rationale. | Included in methods |
| Information sources | 7 | Describe all information sources (e.g., databases with dates of coverage, contact with study authors to identify additional studies) in the search and date last searched. | Included in methods |
| Search | 8 | Present full electronic search strategy for at least one database, including any limits used, such that it could be repeated. | Included in suppl table 2 |
| Study selection | 9 | State the process for selecting studies (i.e., screening, eligibility, included in systematic review, and, if applicable, included in the meta-analysis). | Included in methods |
| Data collection process | 10 | Describe method of data extraction from reports (e.g., piloted forms, independently, in duplicate) and any processes for obtaining and confirming data from investigators. | Included in methods |
| Data items | 11 | List and define all variables for which data were sought (e.g., PICOS, funding sources) and any assumptions and simplifications made. | Included in methods |
| Risk of bias in individual studies | 12 | Describe methods used for assessing risk of bias of individual studies (including specification of whether this was done at the study or outcome level), and how this information is to be used in any data synthesis. | Included in methods (data extraction section) |
| Summary measures | 13 | State the principal summary measures (e.g., risk ratio, difference in means). | Incidence of complications is summary measure – |

| | | | |
|---|---|---|---|
| | | | presented in results and Fig 3 |
| Synthesis of results | 14 | Describe the methods of handling data and combining results of studies, if done, including measures of consistency (e.g., $I^2$) for each meta-analysis. | Fig 3 |

| **Section/topic** | **#** | **Checklist item** | **Reported on page #** |
|---|---|---|---|
| Risk of bias across studies | 15 | Specify any assessment of risk of bias that may affect the cumulative evidence (e.g., publication bias, selective reporting within studies). | Bias in selection criteria – tables S5 and S6 |
| Additional analyses | 16 | Describe methods of additional analyses (e.g., sensitivity or subgroup analyses, meta-regression), if done, indicating which were pre-specified. | No additional methods used |
| **RESULTS** | | | |
| Study selection | 17 | Give numbers of studies screened, assessed for eligibility, and included in the review, with reasons for exclusions at each stage, ideally with a flow diagram. | Presented in results and flow diagram (Fig 2) |
| Study characteristics | 18 | For each study, present characteristics for which data were extracted (e.g., study size, PICOS, follow-up period) and provide the citations. | Summarised in tables S5 and S6 |
| Risk of bias within studies | 19 | Present data on risk of bias of each study and, if available, any outcome level assessment (see item 12). | Final column in tables S5 and S6; specific 'risk of bias' sub-section in results |
| Results of individual studies | 20 | For all outcomes considered (benefits or harms), present, for each study: (a) simple summary data for each intervention group (b) effect estimates and confidence intervals, ideally with a forest plot. | Forest plots presented (Fig 3) |
| Synthesis of results | 21 | Present results of each meta-analysis done, including confidence intervals and measures of consistency. | Formal meta-analysis not done due to insufficient data, but pooled estimates are presented |
| Risk of bias across studies | 22 | Present results of any assessment of risk of bias across studies (see Item 15). | Specific 'risk of bias' sub-section in results, with further comments in discussion section |
| Additional analysis | 23 | Give results of additional analyses, if done (e.g., sensitivity or subgroup analyses, meta-regression [see Item 16]). | No additional methods used |
| **DISCUSSION** | | | |
| Summary of evidence | 24 | Summarize the main findings including the strength of evidence for each main outcome; consider their relevance to key groups (e.g., healthcare providers, users, and policy makers). | Included in discussion |

| | | | |
|---|---|---|---|
| Limitations | 25 | Discuss limitations at study and outcome level (e.g., risk of bias), and at review-level (e.g., incomplete retrieval of identified research, reporting bias). | Included in discussion |
| Conclusions | 26 | Provide a general interpretation of the results in the context of other evidence, and implications for future research. | Included in discussion |
| **FUNDING** | | | |
| Funding | 27 | Describe sources of funding for the systematic review and other support (e.g., supply of data); role of funders for the systematic review. | Funders included at the end of the manuscript |

*From:* Moher D, Liberati A, Tetzlaff J, Altman DG, The PRISMA Group (2009). Preferred Reporting Items for Systematic Reviews and Meta-Analyses: The PRISMA Statement. PLoS Med 6(7): e1000097. doi:10.1371/journal.pmed1000097. For more information, visit: **www.prisma-statement.org**.

**Table S4:** Summary characteristics of observational studies investigating risk of TB infection in individuals receiving tocilizumab. Studies reporting incidence of TB reactivation on a per-patient basis are included in the summary figure in the main text (Fig 3A).

| Country, Author (Year) | Study design | Population | Follow-up period | Participants receiving tocilizumab, n | TB Cases, n | Age at baseline, years | Sex (% male) | Notes and risk of bias |
|---|---|---|---|---|---|---|---|---|
| Hong Kong, Chung (2020) (1) | Retrospective cohort study | Patients with RA or non-specific back pain | 11.3 years(med)* | 49 | 1 | >60 (med) * | <50 * | Previous episodes of TB infection and the sites of TB infection were recorded, but patients not excluded/included based on this. |
| Hong Kong, Tang (2020) (2) | Retrospective cohort study | Rheumatic disease patients receiving DMARDs | >=6 months | 40 | 2 | 53 (mean) * | 35* | Participants all underwent either single (TST or IGRA) or dual (TST and IGRA) test for LTBI before starting biologic treatment |
| Argentina, Cerda (2019) (3) | Retrospective cohort study | Patients with RA, JIA and SpA receiving treatment with anti-TNF, TCZ and/or abatacept | 14.8 months (med)* | 13 | 1 | 52 (mean)* | 21* | Only patients with a previous negative TST test were eligible, and all received a second TST test within 22 months of the first test. Patients with history of active TB or LTBI, or those with two tests performed >22 months apart, were excluded. |
| Taiwan, Lin (2019) (4) | Prospective cohort study | RA patients receiving TCZ | 3 years | 114 | 0 | 59 (med) | 15 | Patients screened for LTBI and prophylactic isoniazid administered in 12 patients |
| Hong Kong, Mok (2019) (5) ‡ | Retrospective cohort study | RA patients ever treated with DMARDs | 3.7 years (mean) * | NA | NA† | 54 (mean) * | 12* | No information on TB screening |
| Taiwan, Huang (2018) (6) | Retrospective cohort study | Patients aged >=20 years with immune-mediated inflammatory disorders | 19 months (med) * | 16 | 3 | NA (80% aged <=65) * | 33* | Participants screened for LTBI with IGRA at baseline, those with positive screen offered isoniazid, which a portion of participants refused |
| India, Malaviya (2018) (7) | Retrospective cohort study | Patients with inflammatory rheumatic musculoskeletal | >= 1 year | 7 | 2 | 32 (med) * | 57* | Patients screened LTBI using augmented Manteau and QFTG test, and positive patients offered prophylaxis |

| Country, Author (Year) | Study design | Population | Follow-up period | Participants receiving tocilizumab, n | TB Cases, n | Age at baseline, years | Sex (% male) | Notes and risk of bias |
|---|---|---|---|---|---|---|---|---|
| | | disease receiving biologic DMARDs | | | | | | |
| UK, Rutherford (2018) (8) | Prospective cohort study | RA patients receiving biologic therapy | 1.8 years (mean) | 2171 | 1 | NA | NA | Patients screened according to clinical guidelines at time of treatment initiation (cohort established in 2001) |
| Italy, Cuomo (2017) (9) | Prospective cohort study | RA patients | 10 years (total study duration)* | 44 | 7 | 55 (mean) * | 16* | All patients required negative screen for LTBI at baseline (screened before starting therapy with TST and QFTG assay). |
| Taiwan, Lim (2017) (10) | Retrospective cohort study | RA patients receiving biologic DMARDs | 1.8 years (mean) | 31 | 0 | 54 (mean) * | 26 | Patients screened for LTBI via QFTG assay |
| France, Devauchelle-Pensec (2016) (11) | Prospective open-label study | PMR patients aged 50-80 receiving TCZ | 24 weeks (total study duration) | 20 | 0 | 67 (med) | 65 | No information on TB screening |
| France, Morel (2016) (12) | Retrospective cohort study | RA patients receiving TCZ | 1.8 years (mean) | 1491 | 1 | 57 (mean) | NA | No information on TB screening |
| Argentina, Benzaquen (2015) (13) | Case-control | RA patients receiving biologic DMARDs age- and sex-matched to RA patients receiving non-biologic DMARDs | 5 years (total study duration)* | 107 | NA† | 43 (mean) * | 21* | No information on TB screening |

| Country, Author (Year) | Study design | Population | Follow-up period | Participants receiving tocilizumab, n | TB Cases, n | Age at baseline, years | Sex (% male) | Notes and risk of bias |
|---|---|---|---|---|---|---|---|---|
| Italy, Cuomo (2015) (14) | Prospective cohort study | RA patients | 1 year (total study duration) * | 28 | 1 | 55 (mean) * | 10* | Patients screened negative for LTBI at baseline via TST and QFTG assay; re-screened at one year |
| France, Gottenberg (2015) (15) | Retrospective cohort study | RA patients treated with abatacept, rituximab or TCZ | 1 year (mean)* | 1503 | 0 | NA | NA | No information on TB screening |
| Japan, Yamamoto (2015) (16) | Extension of a single-arm, observational post-marketing surveillance study | RA patients treated with TCZ | 3 years (total study duration) | 5573 | 8 | 59 (mean) | 18 | Patients screened for TB via chest radiograph, tuberculin test, and interview at baseline, but patients not excluded based on result |
| Hong Kong, Mok (2014) (17) | Retrospective cohort study | Patients with rheumatic disease receiving biologic DMARDs | 1.3 years (mean) | 159 | 0 | NA | 42* | Routine screening for LTBI using TST. Patients were given isoniazid treatment for LTBI before starting therapy. |
| Germany, Burmester (2013) (18) | Prospective cohort study | RA patients receiving TCZ | 1 year (total study duration) | 850 | 2 | 56 (mean) | 25 | No information on TB screening |
| International, Genovese (2013) (19) | Pooled analysis from 5 RCTs and their open-label extension phases | RA patients included in TCZ randomised controlled trials | 3 years (mean)* | 2644 | 9 § | NA | NA | Patients screened for TB and those with active TB were excluded, but those with LTBI received standard of care prophylaxis and were eligible for inclusion |
| South Africa, Hansrajh (2013) (20) | Open-label extension study | RA patients included in TCZ randomised controlled trial | 24 weeks (total observation duration) | 29 | 0 | 56 (med) | 17 | Patients screened for LTBI and prophylaxis provided |
| Japan, Koike (2011) (21) | Prospective post-marketing | RA patients treated with TCZ | 28 weeks (total | 3881 | 4 | NA | NA | Patients screened for TB via chest radiograph, tuberculin test, and interview before initiation of TCZ |

| Country, Author (Year) | Study design | Population | Follow-up period | Participants receiving tocilizumab, n | TB Cases, n | Age at baseline, years | Sex (% male) | Notes and risk of bias |
|---|---|---|---|---|---|---|---|---|
| | surveillance study | | observation duration) | | | | | |

TB, tuberculosis; RA, rheumatoid arthritis; TCZ, tocilizumab; DMARD, disease-modifying anti-rheumatic drug; TST, tuberculin skin test; IGRA, interferon gamma releasing assay; LTBI, latent tuberculosis infection; JIA, juvenile idiopathic arthritis; SpA, spondyloarthritis; TNF, tumour necrosis factor; QFTG, QuantiFERON®-TB Gold; AS, ankylosing spondylitis; PsA, psoriatic arthritis; PMR, polymyalgia rheumatica.

* Value for whole cohort; value for subset receiving TCZ not reported.

† Authors only report rate of TB infection: 0.36 cases/100 patient-years

‡Conference/published abstract

§ 8 cases thought to be *de novo*

**Table S5:** Summary characteristics of observational studies investigating risk of HBV reactivation in individuals receiving tocilizumab.

| Country, Author (Year) | Study design | Population | Follow-up period | Participants, n | HBVr Cases, n | Age at baseline, years | Sex (% male) | Notes and risk of bias |
|---|---|---|---|---|---|---|---|---|
| Taiwan, Kuo (2021) (22) | Retrospective cohort study | RA patients receiving TCZ | 9 (med) | 97 | 4 | 64 (med) | 23 | Patients were only eligible if they had HBsAg and anti-HBc at baseline. 7 patients were HBsAg+ve. Out of 90 HBsAg-ve 64 were anti-HBc+ve. 3 HBsAg-positive patients who did not receive prophylaxis all experienced reactivation. One anti-HBs-ve patient experienced reactivation |
| Spain, Rodríguez-Tajes (2021) (23) | Prospective cohort study | Patients with COVID-19 | 1-1.5 months | 44 | 1 | 67* | 72* | HBsAg+ve patients excluded. Antiviral prophylaxis strongly recommended in all antiHBc+ve patients: 38 out of 69 patients in the cohort received prophylaxis |
| Taiwan, Lin (2019) (24) | Prospective cohort study | RA patients receiving TCZ | 3 | 114 | 0 | 59 (med) | 15 | Patients screened for HBV and prophylactic NA therapy administered in 11 CHB patients (in whom HBsAg and anti-HBc status was not reported) |
| Japan, Watanabe (2019) (25) | Retrospective cohort study | RA patients treated with biologic DMARDs | 15 months* | 25 | 1 | 68* | 21* | Patients who were anti-HBc and HBsAg-ve and HBV DNA-ve before DMARD initiation were eligible |
| Korea, Ahn (2018) (26) | Retrospective cohort study | RA patients treated with TCZ | NA (>3 months) | 39 | 0 | 55 (med) | 28 | All had HBV serology at baseline. All were HBsAg-negative, and 15 patients had resolved HBV infection (HBsAg-negative; anti-HBc positive). |
| China, Chen (2017) (27) | Prospective cohort study | RA patients treated with TCZ, with moderate to high disease activity and at least one | 12-80 weeks | 48 | 3 | 46* | 21* | Seven patients had CHB and 41 had resolved HBV infection. All cases or HBVr were in CHB (HBsAg+ve) patients. |

| Country, Author (Year) | Study design | Population | Follow-up period | Participants, n | HBVr Cases, n | Age at baseline, years | Sex (% male) | Notes and risk of bias |
|---|---|---|---|---|---|---|---|---|
| | | feature of poor prognosis | | | | | | |
| Hong Kong, Mok (2014) (17) | Retrospective cohort study | Patients with rheumatic disease receiving biologic DMARDs | 1.3 years | 159 | 0 | NA | 42* | No information for HBV screening |
| Japan, Nakamura (2014) (28) | Retrospective cohort study | RA patients treated with biologic DMARDs | 18 months* | 18 | 2 | >=60* | 17* | Mix of patients who were HBsAg+ve/-ve and anti-HBc+ve/-ve at baseline. No patients received prophylactic antiviral therapy. |

RA, rheumatoid arthritis; HBVr, hepatitis B virus reactivation; HBsAg, hepatitis B serum antigen; HBcAb, hepatitis B core antibody; +ve, positive; -ve, negative; TCZ, tociizumab; CHB, chronic hepatitis B; HBc, hepatitis b virus core antigen; DMARD, disease-modifying anti-rheumatic drugs; DNA, deoxyribonucleic acid; TST, tuberculin skin test.

*Value for whole cohort; value for subset receiving TCZ not reported.

**Table S6:** Summary of papers reporting an influence of tocilizumab on HCV infection

| Author, date of publication | Type of study | Conclusions and/or clinical recommendation(s) |
|---|---|---|
| Biehl et al., 2021 (29) | Case series | Flare of HCV responsible for liver injury in 1 of 12 cases of liver injury related to tocilizumab |
| Sebastiani et al., 2019 (30) | Review and expert consensus recommendations | Recommendations that screening for HCV should be performed in all rheumatoid patients before starting immunosuppressive therapy. |
| Chiu et al., 2020 (31) | Systematic review | tbc |
| Banerjee et al., 2021 (32) | Network theoretic analysis | Theoretical analysis of the impact of immunosuppressive agents on the network size of the JAK-STAT pathway |
| Karadag et al., 2016 (33) | Clinical guidelines | Recommendations for any individual with risk factors for HCV infection to be screened prior to biologic therapy, but treatment generally does not cause deterioration (exception is rituximab) |
| Giannitti et al., 2013 (34) | Case report | TCZ + cyclosporin A treatment for rheumatoid in a patient with HCV was tolerated without any flare in liver enzymes |
| Song et al., 2019 (35) | Mechanistic study | Investigated the association of IL6 (via the signal transducers and activators of transcription 3 (STAT3) pathway) with HCV infection, but no specific assessment of the impact of TCZ |

**Table S7:** Summary of review articles presenting data on the risks of HBV, HCV and/or TB reactivation in patients on TCZ therapy, identified by systematic literature review

| FIrst author and title of paper | Approach | Conclusions and clinical recommendation(s) |
|---|---|---|
| Alqahtani, et al., 2021, COVID-19 and hepatitis B infection (36) | Narrative summary of interaction between COVID and HBV infection | • Highlights need for caution regarding potential for HBV reactivation following treatment with TCZ and corticosteroids.<br>• Recommends HBV screening/prophylaxis for patients with elevated transaminases and also in high prevalence populations. |
| Boyman et al., 2014, Adverse reactions to biological agents and their medical management (37) | Narrative summary of adverse effects of biologic therapy used for treatment of chronic inflammatory and autoimmune disorders | • Reports association between TCZ and increased risk of serious infections, including increased risk of reactivation of latent TB; however, synthesis of literature suggests no overall increase in risk of SIE in patients on TCZ<br>• Recommends TB screening, and subsequent prophylaxis based on risk assessment.<br>• Highest risk of severe infections in the first weeks-months of treatment with biologic agents.<br>• Patients with abnormal LFTs should undergo HBV testing before use of TCZ. |
| Craig et al., 2018, Gastrointestinal and Hepatic Disease in Rheumatoid Arthritis (38) | Narrative summary of GI complications of Rheumatoid and its treatment | • HBV reactivation recognised as a complication of corticosteroids, but not specifically listed in association with TCZ.<br>• Reactivation of TB recognised, with a higher rate of extrapulmonary disease than in patients not receiving biologic therapy.<br>• No specific recommendations about screening or prophylaxis. |
| Felis-Giemza et al., 2015, Treatment of rheumatic diseases and hepatitis B virus coinfection (39) | Narrative summary of the HBV disease cycle and points at which biologic disease-modifying anti-rheumatic drugs may influence this cycle | • Highlights exclusion of HBV-infected individuals from most TCZ trials<br>• Recommends consideration of antiviral treatment before TCZ administration in patients with HBsAg seropositivity |
| Ferro et al., 2017, One year in review 2017: novelties in the treatment of rheumatoid arthritis (40). | Summary of studies published in previous calendar year concerning rheumatoid arthritis treatment novelties | • States that in rheumatoid arthritis patients with high risk of infection or latent TB infection, TCZ is a safer treatment choice compared to other anti-rheumatic drugs |

| | | |
|---|---|---|
| Lai et al., 2019, Useful message in choosing optimal biological agents for patients with autoimmune arthritis (41) | Comparison between biologic agents used alone or in combination for inflammatory / autoimmune arthritis | • TCZ assigned lower risk of TB reactivation compared to anti-TNF therapy for Rheumatoid.<br>• Risk of TB and HBV reactivation recognised with TCZ therapy; TCZ classed as 'intermediate' risk compared to other biologic therapy. |
| Lunel-Fabiani et al., 2014, Systemic diseases and biotherapies: Understanding, evaluating, and preventing the risk of hepatitis B reactivation (42) | Summary of guidelines for the prevention and management of HBV in patients prescribed biological therapy | • Treatment recommended for all active HBV carriers meeting criteria for therapy<br>• Antiviral prophylaxis for HBsAg carriers who do not meet criteria for therapy (6-12 months)<br>• Inadequate data in many cases to inform strategy for HBsAg negative / anti-HBc positive cases, though prophylaxis is recommended by some guidelines<br>• Vaccination of patients with negative HBV markers is universally recommended |
| Novosad et al., 2014, Beyond Tumor Necrosis Factor Inhibition: The Expanding Pipeline of Biologic Therapies for Inflammatory Diseases and Their Associated Infectious Sequelae (43) | Summary of data regarding infective complications of biologic therapy | • TB screening is recommended before starting any biologic therapy, with prophylactic therapy in those identified with latent TB.<br>• Screening for HBV and HCV infection recommended before any biologic therapy.<br>• For HBV, give antiviral prophylaxis to those who test HBsAg positive, and close monitoring for HBsAg negative / anti-HBc positive patients with liver function and viral loads |
| Rose-John et al., 2017 The role of IL-6 in host defence against infections: immunobiology and clinical Implications (44) | Review of biology of IL-6 together with pooled analysis of serious infection events in patients treated with different biological agents | • No specific clinical recommendations are stated, but the paper highlights risks of both HBV and TB reactivation supported by data from clinical trials and pooled analyses. |
| Sebastiani et al., 2019, Italian consensus recommendations for the management of hepatitis C infection in patients with rheumatoid arthritis (30). | Consensus process produced to review available evidence and produce recommendations regarding HCV management in patients with rheumatoid arthritis | • Screening for HCV should be mandatory before initiation of rheumatoid treatment, and antiviral treatment should be considered in all HCV-infected patients.<br>• HCV eradication should be pursued in all HCV-infected patients, with eradication attempted before initiation of treatment.<br>• Lack of evidence regarding TCZ safety in HCV-infected patients highlighted. |

| | | |
|---|---|---|
| Winthrop et al., 2018 ESCMID Study Group for Infections in Compromised Hosts (ESGICH) Consensus Document on the safety of targeted and biological therapies (45) | Consensus statement based on literature review, summarised into recommendations by expert opinion | • Infection risks associated with TCZ are quantified as similar to anti-TNF therapy.<br>• Screening for HBV should be undertaken prior to starting therapy<br>• Antiviral prophylaxis should be prescribed for HBsAg-positive patients.<br>• Screening should be undertaken for latent TB infection, followed by appropriate prophylaxis or treatment. |

anti-HBc - antibody to hepatitis B virus core antigen; ESCMID - European Society of Clinical Microbiology and Infectious Diseases; HBV - hepatitis B virus; HBsAg - hepatitis B surface antigen; HCV - hepatitis C virus; SIE - severe infection events; TB - tuberculosis; TCZ - tocilizumab; TNF - tumour necrosis factor
